# The PREVENT-AD cohort: accelerating Alzheimer’s disease research and treatment in Canada and beyond

**DOI:** 10.1101/2025.07.22.25331791

**Authors:** Sylvia Villeneuve, Judes Poirier, John C.S. Breitner, Jennifer Tremblay-Mercier, Jordana Remz, Jean-Michel Raoult, Yara Yakoub, Jonathan Gallego-Rudolf, Ting Qiu, Alfonso Fajardo Valdez, Bery Mohammediyan, Mohammadali Javanray, Amelie Metz, Safa Sanami, Valentin Ourry, Alfie Wearn, Alexandre Pastor-Bernier, Manon Edde, Julie Gonneaud, Cherie Strikwerda-Brown, Christine L. Tardif, Claudine J. Gauthier, Maxime Descoteaux, Mahsa Dadar, Étienne Vachon- Presseau, Andree-Ann Baril, Simon Ducharme, Maxime Montembeault, Maiya R. Geddes, Jean-Paul Soucy, Natasha Rajah, Robert Laforce, Christian Bocti, Christos Davatzikos, Lune Bellec, Pedro Rosa-Neto, Sylvain Baillet, Alan C. Evans, D. Louis Collins, M. Mallar Chakravarty, Kaj Blennow, Henrik Zetterberg, R. Nathan Spreng, Alexa Pichet Binette, the PREVENT-AD Research Group

## Abstract

The PREVENT-AD is an investigator-driven study that was created in 2011 and enrolled cognitively normal older adults with a family history of sporadic AD. Participants are deeply phenotyped and have now been followed annually for more than 12 years [median follow-up 8.0 years,SD 3.1]. Multimodal MRI, genetic, neurosensory, clinical, cerebrospinal fluid and cognitive data collected until 2017 on 348 participants who agreed to open sharing with the neuroscience community were already available. We now share a new release including 6 years of additional follow-up cognitive data, and additional MRI follow-ups, clinical progression, new longitudinal behavioral and lifestyle measures (questionnaires, actigraphy), longitudinal AD plasma biomarkers, amyloid-beta and tau PET, magnetoencephalography, as well as neuroimaging analytic measures from all MRI modalities. We describe the PREVENT-AD study, the data shared with the global research community as well as the model we created to sustain longitudinal follow-ups while also allowing new innovative data collection.

## 1. HISTORY, GOVERNANCE AND DATA SHARING MODELS

The Centre for Studies on the Prevention of Alzheimer’s Disease (StoP-AD Centre, www.centre-stopad.com/) was created in 2010, at the Douglas Mental Health University Institute Research Centre, under the leadership of Dr. John C.S. Breitner, MD, MPH, Dr. Judes Poirier, PhD, and Dr. Pierre Etienne, MD as a 7-year $13.5M public-private partnership supported by McGill University^1^. The current director of the Centre is Dr. Sylvia Villeneuve, PhD, and Dr. Judes Poirier is Deputy Director. The main mission of the StoP-AD Centre is the pursuit of innovative intersectoral studies and interventions in the preclinical, or asymptomatic, phase of Alzheimer’s disease (AD), with the goal of stopping or delaying the onset of the disease.

The primary resource of the StoP-AD Centre is the PResymptomatic EValuation of Experimental or Novel Treatments for Alzheimer’s disease (PREVENT-AD) cohort, which includes individuals cognitively unimpaired (CU) at the time of enrolment with a first-degree family history of sporadic Alzheimer-like dementia. Having a family history of sporadic AD is known to increase the risk of dementia by 2- to 3-fold^2^. These “at-risk” individuals undergo annual detailed multimodal examinations across a diverse array of disease indicators, such as neuropsychological evaluations, biofluid collection, and neuroimaging markers.

To build the PREVENT-AD cohort, more than 1700 persons from the greater Montreal area were screened between 2011 and 2017. Of these, 692 completed an on-site eligibility visit, 426 completed a baseline visit and 387 were included in the cohort (**Figure 1**)^3^. Participants needed to have a parental history of sporadic Alzheimer-like dementia or multiple siblings affected by the disease and be aged 60 years or older, or 55–59 years if within 15 years of the age of onset of their affected relative. At the eligibility visit, the Montreal Cognitive Assessment (MoCA) and the Clinical Dementia Rating (CDR) were administered to exclude the presence of cognitive impairment. A very small portion of individuals who had a MoCA lower than 26 or a CDR higher than 0 were enrolled after an exhaustive neuropsychological evaluation performed by a clinician that confirmed normal cognition. The Repeatable Battery for the Assessment of Neuropsychological Status (RBANS)^4^ was performed at baseline, and participants with scores below normative values also needed to undergo an exhaustive neuropsychological evaluation to confirm normal cognition otherwise they were excluded from the study. The full list of inclusion/exclusion criteria can be found in **supplementary Table 1**^1,3^. Among the 387 participants included in the PREVENT-AD study, 367 completed at least one follow-up visit (95%) and 306 (79%) were still followed more than 10 years later.

**Figure 1:**
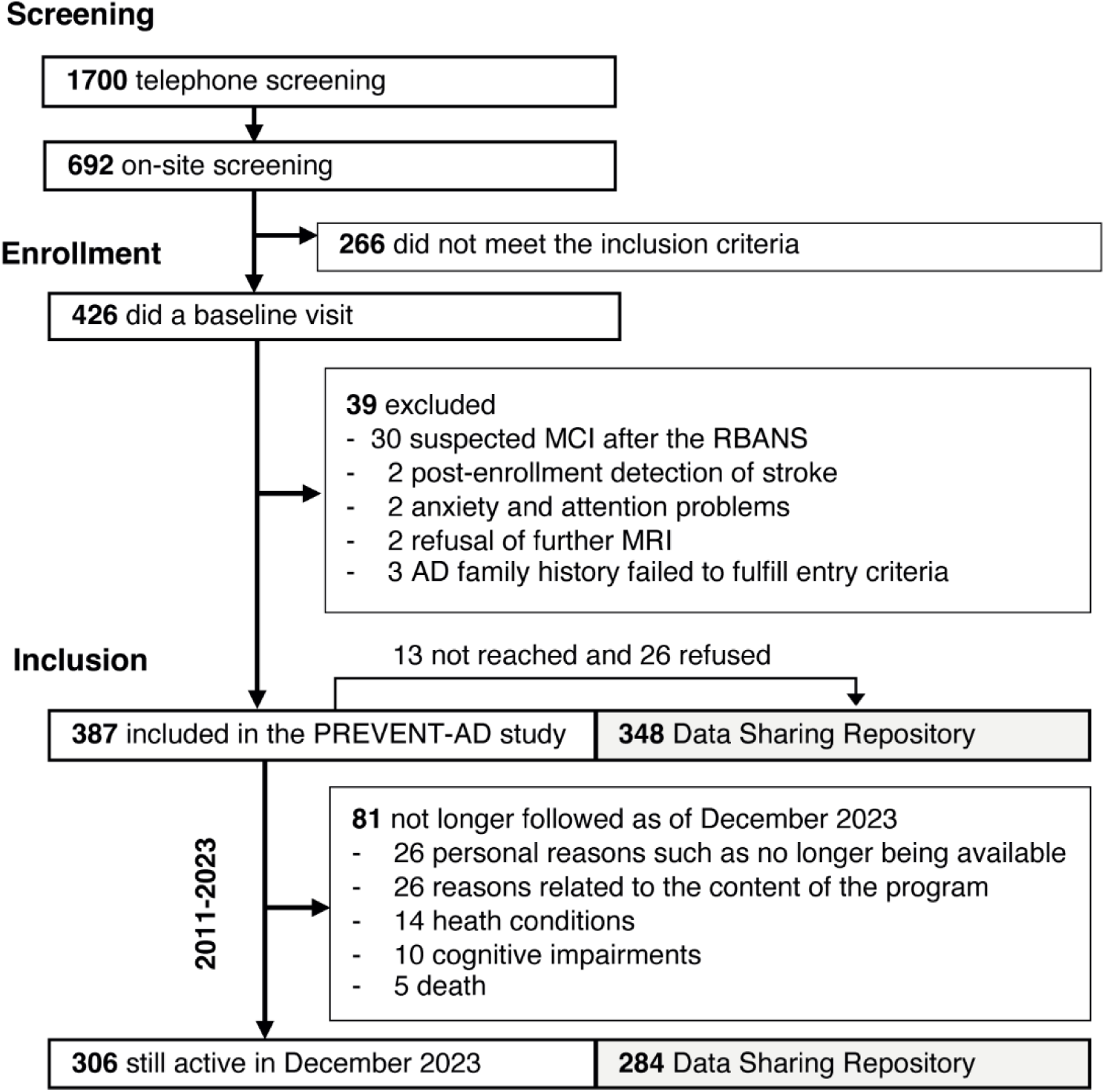
Flow chart of PREVENT-AD parti 1.

The mean age at baseline of the participants was 63 years (SD=5.2 years), 72% were female, 38% were *APOE*4 carriers, and 98% were White (**supplementary Table 2).** 87% had French as a mother tongue and 74% reported being bilingual or multilingual. Between 2011-2017, corresponding to Phase 1 of data acquisition, participants were seen annually and data collected included MRI, cognitive, neurosensory (olfactory and auditory processes) and biofluid (plasma, urine and cerebrospinal fluid [CSF])^3^. Two pharmacological trials were also completed. A 2-year double-masked pharmaco-prevention trial (naproxen vs placebo), that enrolled 195 of the 387 PREVENT-AD participants was conducted between November 2011 and March 2017 (NCT02702817)^5^. The trial provided no conclusive evidence for superiority of naproxen over placebo. A second trial targeting apolipoprotein E (APOE) production was started in 2016, but rapidly terminated as it did not reach its short-term goals (NCT02707458). Lumbar punctures (LP) in the PREVENT-AD cohort were optional and initially proposed only to participants enrolled in the naproxen trial, before being proposed to the full cohort in 2016.

In November 2017, the McGill University funding period concluded, forcing a restructuring of the StoP-AD Centre’s operations and governance. At that time, 341 participants were still followed. We moved from a top-down data collection structure to a bottom-up structure where collaborating investigators could propose innovative projects to be conducted with PREVENT-AD participants while also supporting acquisition of the “core” data collected in the Phase 1 (e.g. RBANS), to balance continuity and innovation. The proposed projects were carefully reviewed to ensure they were in line with the Centre’s main objectives and did not overburden the participants.

During this restructuring, we also received support from the Canadian Open Neuroscience Platform (CONP) to share PREVENT-AD data with the global research community. When initially enrolled in the cohort, participants consented to their data being used by StoP-AD investigators and collaborators only. We therefore retrospectively consented participants for their data to be shared with neuroscience researchers worldwide. We were able to reach 373 of the 387 PREVENT-AD participants among which 348 (93% of those reached) agreed to such sharing. The first open release of PREVENT-AD data as well as the steps needed to move from a traditional data sharing model to a global sharing model have been described in detail previously^3^.

The overarching goal of this manuscript is to describe the main data available to PREVENT-AD investigators and to researchers worldwide, with a focus on the data collected in the second phase of data acquisition (overview in **Figure 2**). This second phase of data acquisition includes 6 years of additional follow-up cognitive data, and additional MRI follow-ups, clinical progression, new longitudinal behavioral and lifestyle measures (questionnaires, actigraphy), longitudinal AD plasma biomarkers, amyloid-beta (Aβ) and tau PET, magnetoencephalography, as well as neuroimaging analytic measures from all MRI modalities. **Figure 3A** provides a timeline of the main in-person data collected on PREVENT-AD participants at the StoP-AD Centre, and **Figure 3B** summarizes the data collected across baseline and follow-up visits for all participants. The data of the 348 participants who consented to global sharing can be accessed by faculty researchers and physicians at https://registeredpreventad.loris.ca after accepting the terms of use, such as not trying to re-identify participants.

**Figure 2:**
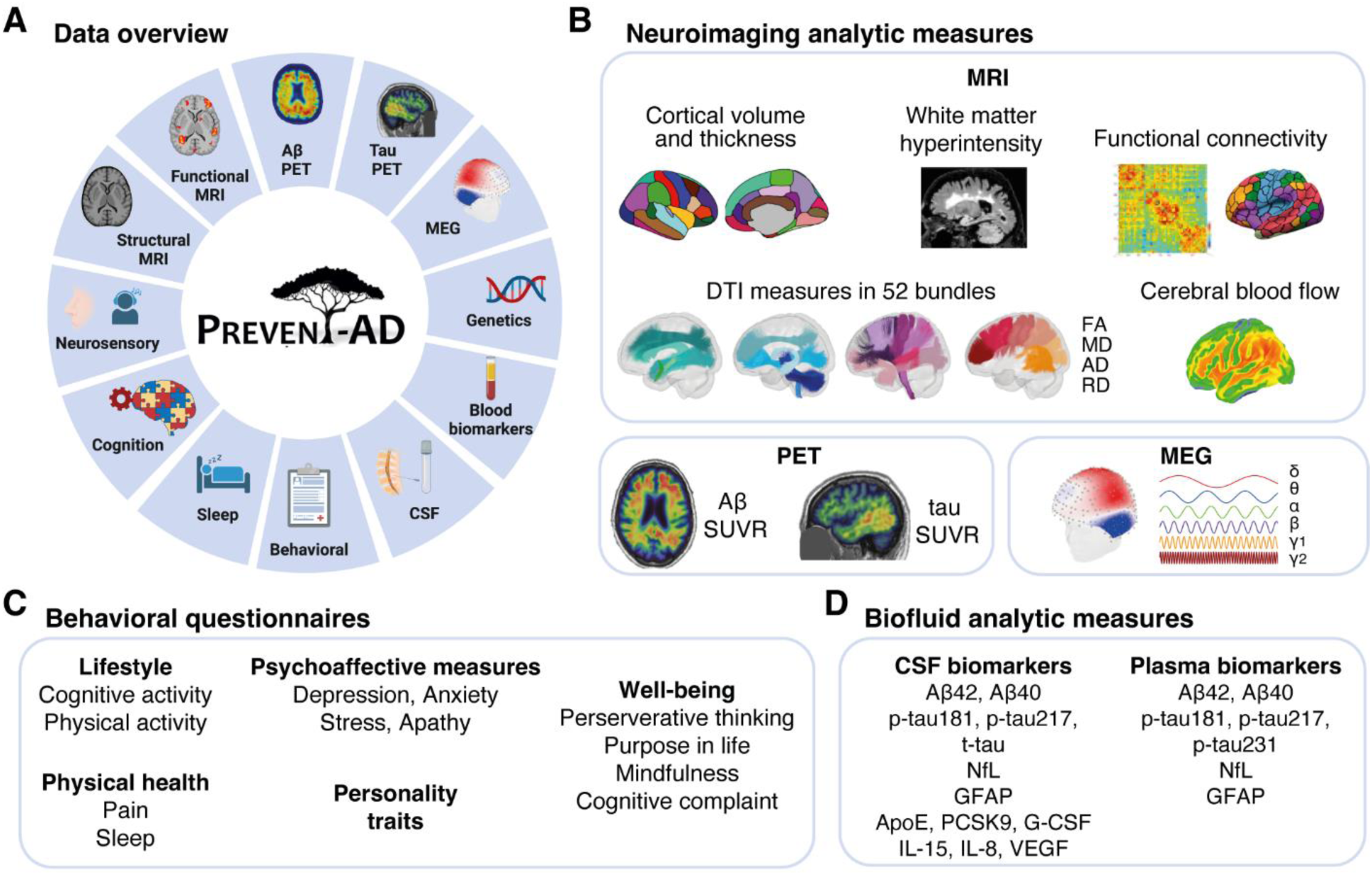
Main data available in the Data Shearing Repository. Overview of all modalities available (**A**). Neuroimaging analytic measures (**B**). The cognitive assessment includes global and specific neuropsychological tests. Behavioral questionnaires (**C**). Fluid analytic measures (**D**). *Abbreviations*: Aβ, Amyloid-beta; PET, positron emission tomography; MEG, magnetoencephalography; CSF, cerebrospinal fluid; MRI, magnetic resonance imaging; PD, proton density; MTsat, magnetization transfer saturation; DTI, diffusion tensor imaging; FA, fractional anisotropy; MD, mean diffusivity; AD, axial diffusivity; RD, radial diffusivity; SUVR, standardized uptake value ratio; p, phosphorylated; NfL, neurofilament light; GFAP, glial fibrillary acidic protein; *APOE*, Apolipoprotein E; PCSK9, proprotein convertase subtilisin/kexin type 9; G-CSF, granulocyte colony stimulating factor; IL, interleukin; VEGF, vascular endothelial growth factor.

**Figure 3:**
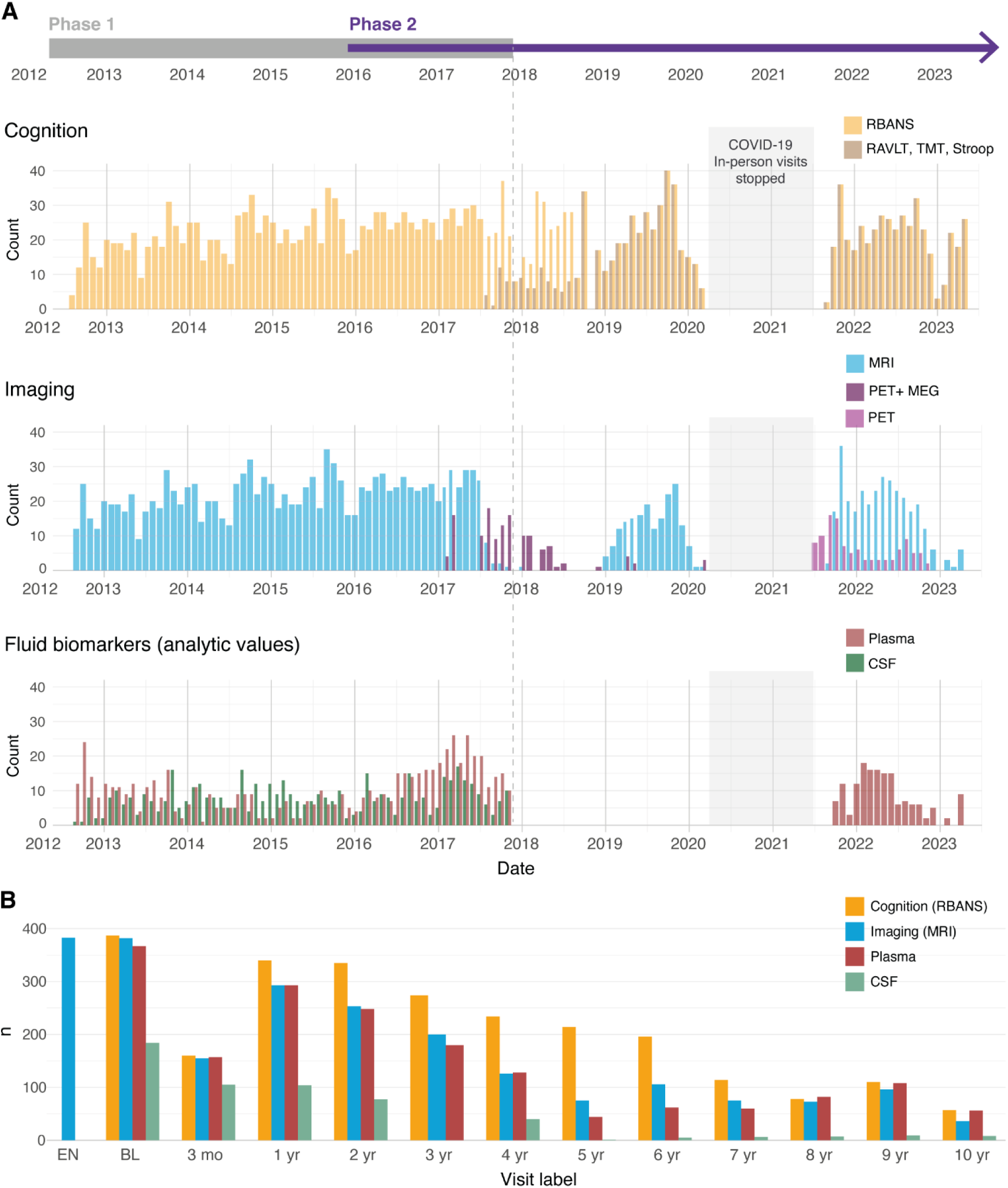
Main data collected over the years in the PREVENT-AD cohort. Overview of the in-person data collected with PREVENT-AD participants at the StoP-AD Centre between 2012 and 2023 (**A**). The end of Phase 1 is identified by a dotted line. Summary of the baseline and follow-up visits performed on the 387 participants (**B**). The RBANS data collected between March 2020 and July 2021 were obtained virtually due to the COVID 19 pandemic and are not shared given that all tasks have not been completed, and we could not compute index scores. 2011 is not shown given that only enrolment visits were conducted that year. *Abbreviations:* RBANS, Repeatable Battery for the Assessment of Neuropsychological status; RAVLT, Rey Auditory Verbal Learning Test; TMT, Trail Making Test; MRI, magnetic resonance imaging; PET, positron emission tomography; MEG, magnetoencephalography; CSF, cerebrospinal fluid; EN, enrolment; BL, baseline.

## 2. PHASES OF DATA ACQUISITION

### Phase 1

Demographic, anthropometric and genetic (including *APOE* genotype) information were collected at baseline. Baseline and longitudinal cognition were evaluated using the RBANS, which consists of 12 subtests (list learning, story learning, figure copy, line orientation, picture naming, semantic fluency, digit span, coding, list recall, list recognition, story recall, figure recall) that yield an index score for each of five specific cognitive domains (immediate memory, delayed memory, language, attention and visuospatial capacities) and a global score. The Eight-item Informant Interview to Differentiate Aging and Dementia (AD8) was assessed at baseline and annual visits up to November 2017 to evaluate possible mild dementia. Biofluid (blood, urine at each visit; CSF was optional) and sensory (olfactory and auditory) information were also collected. Multi-modal MRI was performed on an annual basis. Most MRI sequences (3T) were harmonized with the Alzheimer’s Disease Neuroimaging Initiative (ADNI) protocol^6^ to ease interoperability between cohorts and included T1-weighted, T2*-weighted, fluid-attenuated inversion recovery (FLAIR) images, diffusion MRI, arterial spin labeling (ASL) and resting-state functional (f)MRI data acquired with a Siemens Tim Trio with a standard 12-channel coil (Siemens Medical Solutions, Erlangen, Germany)^7–14^. An episodic memory fMRI task (designed by Dr. Natasha Rajah, PhD) was also performed for all participants except the ones enrolled after June 2016^15–18^. For participants enrolled after June 2016 until November 2017, the task fMRI was replaced by an MP2RAGE for quantitative T1 maps^19^, a multi-echo gradient echo for T2* maps, and a high in-plane resolution T2-weighted image to assess brain microstructure and segment hippocampal subfields, respectively, and a 32-channel coil was used **(Table 1 and supplementary Table 3).** Finally, except when enrolled in the Naproxen clinical trial, participants who developed mild cognitive impairment (MCI) during the follow-up visits were excluded from the study. Ten participants were excluded between 2012 and 2017.

**Table 1.** Neuroimaging raw data and analytical-derived measures in PREVENT-AD covering November 2011 to December 2023.

| Image type | Participants<br>Internal<br>(Shared) | Visits<br>Internal<br>(Shared) | Investigator<br>group<br>(Acquisition) | Analytical neuroimaging-derived measures | Investigator<br>group<br>(Analytic measures) |
| --- | --- | --- | --- | --- | --- |
| **Magnetic resonance imaging** |  |  |  |  |  |
| T1-weighted<br>(MPRAGE) | 386 (347) | 2264 (2081) | StoP-AD Centre | Regional cortical volume, thickness, surface area, and subcortical volume, quantified using FreeSurfer<br>Machine learning derived measures: brain parcellations and indices of aging and AD using NiChart | S. Villeneuve<br>C. Davatzikos |
| T2-weighted | 304 (286) | 597 (565) | StoP-AD Centre | WMH volume quantified using a random forests machine learning algorithm | M. Dadar |
| Diffusion MRI | 385 (346) | 1465 (1302) | StoP-AD Centre | Diffusion properties of white matter tracts quantified using TractoFlow, RBXFlow and Tractometry Flow | M. Descoteaux |
| Resting-state fMRI | 385 (346) | 1874 (1674) | StoP-AD Centre | Connectome matrices of functional parcellations (post-processing of fMRIPrep outputs) | S. Villeneuve |
| FLAIR | 383 (344) | 1056 (972) | StoP-AD Centre | WMH volume quantified using a random forests machine learning algorithm | M. Dadar |
| Task fMRI | 331 (294) | 1422 (991) | StoP-AD Centre | N/A (Onset vector files available) | N. Rajah |
| T2*-weighted | 381 (343) | 1324 (1261) | StoP-AD Centre | N/A | N/A |
| Arterial spin labeling | 381 (343) | 1243 (1137) | StoP-AD Centre | Cerebral blood flow quantification using FSL | C. Gauthier |
| **Positron emission tomography** |  |  |  |  |  |
| [<sup>18</sup>F]NAV4694<br>(Aβ) | 243 (232) | 243 (232) | S. Villeneuve | Standardized uptake value ratio (SUVR) in DK atlas regions using the whole cerebellum and the cerebellum grey matter as reference regions | S. Villeneuve |
| [<sup>18</sup>F]flortaucipir<br>(tau) | 240 (229) | 240 (229) | S. Villeneuve | SUVR in DK atlas regions using the inferior grey cerebellum as reference region | S. Villeneuve |
| **Magnetoencephalography** |  |  |  |  |  |
| Resting-state MEG | 124 (114) | 124 (114) | S. Villeneuve | Relative spectral power density estimates of delta, theta, alpha, beta, gamma1 and gamma2 frequency bands across cortical regions of the DK atlas | S. Baillet |

*Abbreviations:* DK, Desikan-Killiany; FLAIR, Fluid-attenuated inversion recovery; f, functional; MPRAGE, Magnetization prepared rapid gradient echo; SUVR, standardized uptake value ratio; WMH, white matter hyperintensity.

### Phase 2

The RBANS was (and is) still collected every year, but other assessments became dependent on investigator-driven project grant funds. As such, MRI scans, blood draws and LPs were done, but not necessarily performed on all participants and/or systematically every year. Furthermore, when we resumed MRI scanning in 2019 the 3T Siemens Tim Trio MRI scanner was upgraded to a Prisma Fit and our MRI protocol was updated. Amidst the protocol updates, the fMRI changed from single-to multi-echo, the diffusion MRI from single shell to multi-shell (**supplementary Table 4**).

New investigator-driven data types also began to be collected towards the end of 2016. The novel measures collected in the second phase of data acquisition include 1) additional clinical and experimental cognitive tasks, 2) behavioral data including self-reported questionnaires and objective sleep measures, 3) new MRI sequences, 4) amyloid-beta (Aβ) and tau positron emission tomography (PET) and 5) and magnetoencephalography (MEG) data. Since 2016 we also now systematically review the cognitive performance of all participants and participants who develop MCI or dementia are no longer excluded. As of December 2023, 105 (27%) of the 387 PREVENT-AD participants were classified as having MCI at their latest follow-up time points among which some later received a diagnosis of dementia.

The second phase of data collection and sharing was supported by 15 project grants, two platform grants, philanthropic donations and a Canada Foundation for Innovation award, see the full list in the Acknowledgments section.

## 3. DATA

### Cognitive data

A total of 2662 RBANS assessments have been collected at the StoP-AD Centre between 2012 and 2023 and 2493 of these assessments, representing the visits of the 348 participants who retrospectively agreed to data sharing, are now available to the global research community via the Data Sharing Repository (https://registeredpreventad.loris.ca). RBANS was completed at all visits for all participants except during the COVID-19 pandemic, and most participants have now been followed for 8 years (median follow-up 8.0 years, mean 7.1, standard deviation 3.1, range 0-11.3 years). Starting in August 2017, three new neuropsychological tests were added: the Rey Auditory Verbal Learning Test (RAVLT), Trail Making Test (TMT) and Color-Word Interference Test of the Delis-Kaplan Executive Function System (D-KEFS) on a subset of participants and then on the full cohort. Further details on each of these tests are summarized in **Table 2**.

**Table 2.**
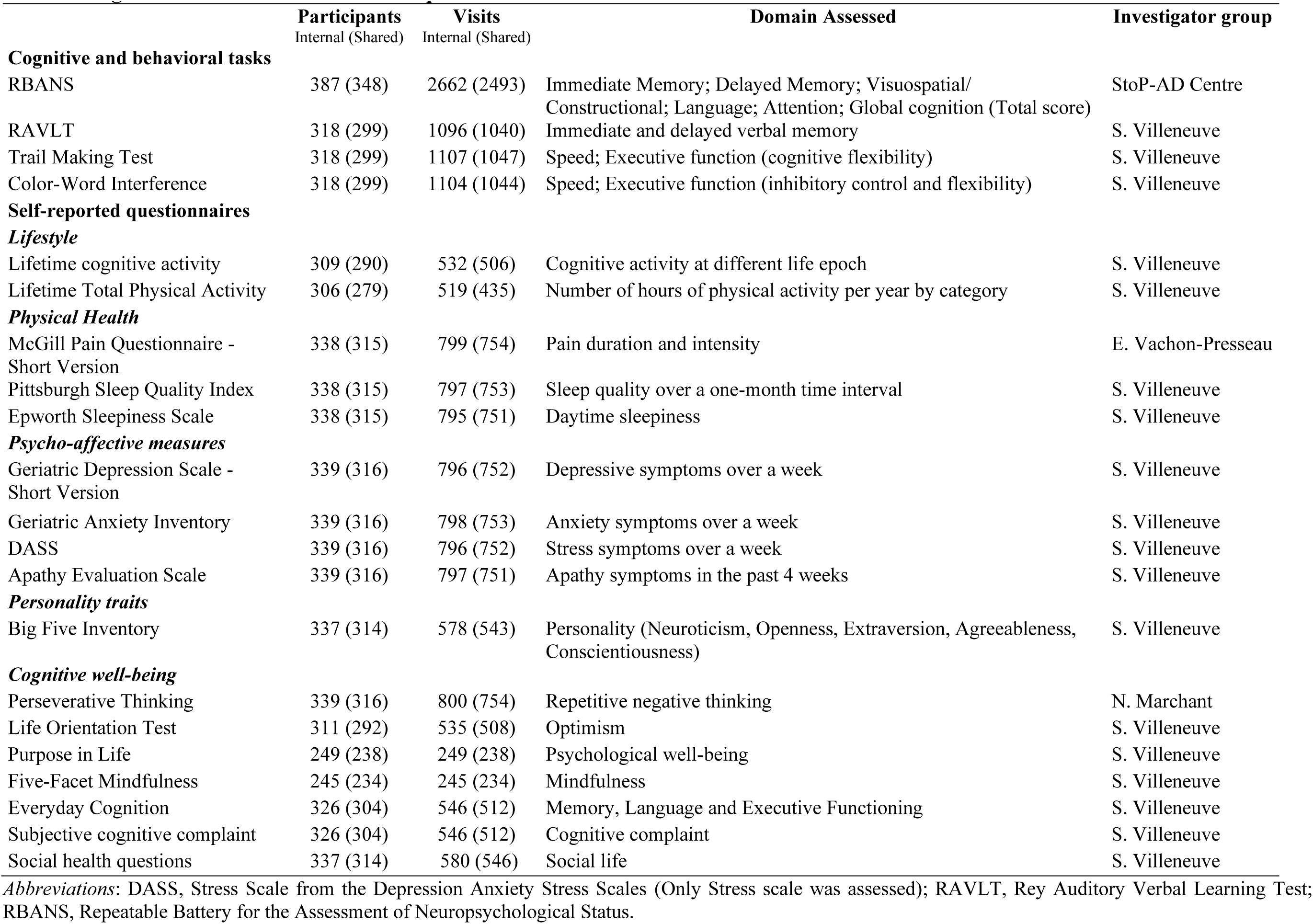
Cognitive and behavioral tasks and questionnaires.

### Clinical progression

To evaluate clinical progression, we reviewed the cognitive evaluations, blind to genetics, imaging and biomarker information, of participants who had a main cognitive complaint or who performed below 1.5 standard deviations in one of the RBANS domains or two sub-tests of the RAVLT^20^. First, trained research staff identified all participants with a main cognitive complaint or who performed below the norms on the RBANS and/or the RAVLT. Then, longitudinal cognitive information of these participants, including all available cognitive time points, were reviewed by two neuropsychologists. Participants who were suspected to have developed MCI based on the neuropsychologist review were then discussed in multidisciplinary consensus meetings with a cognitive neurologist, neuropsychiatrist and the two neuropsychologists. The classification of CU vs MCI was based on all available time points blind to blind to genetics, imaging and biomarker information. The MCI subtype (amnestic or non-amnestic, single or multidomain) was also determined during the consensus meetings. Once a participant was classified as MCI, this label was confirmed at every subsequent visit, meaning that some participants classified as MCI at one visit might revert to be considered as CU at a subsequent visit. Importantly, because the MCI classification was made blind to biomarker information, individuals classified as MCI do not necessarily have AD. Furthermore, most of the participants (66%) with MCI still had a CDR of 0, suggesting that our MCI classification includes participants with milder deficits than what is seen in some other large cohorts, and might represent participants who could be classified as having subjective cognitive decline in other cohorts or if seen in a memory clinic. Finally, some participants can no longer come to their annual visit given the severity of their cognitive impairments and some have received a formal diagnosis of Alzheimer dementia (reported by the study partner or a physician). The main data provided classifies individuals as CU vs MCI based on research classification. An additional CSV file reporting the CDR score, when available, as well as the latest clinical status of the participants, based on non-research information (clinical diagnosis when available or information from caregiver for dementia status) not necessarily blind to biomarker status is also available. 110 participants are classified as having MCI at a minimum of one time point among which 102 are included in the Data Sharing Repository.

### Behavioral data

Personality, lifestyle, mood, and behavioral questionnaires were introduced in 2016 as part of an investigator-led project with the aim of evaluating the impact of modifiable risk factors on AD pathology and cognitive decline^21–27^. The questionnaires cover aspects related to cognitive and physical activity, neuropsychiatric symptoms, personality traits, pain, outlook on life and subjective cognitive complaints (**Table 2**). These questionnaires were mainly collected using online platforms and were not necessarily obtained at the same time as the other assessments.

Questionnaires are available on most participants and have been administered at several time points. Subjective and objective sleep measures were also recorded at multiple follow-ups. Self-reported questionnaires included the Pittsburgh Sleep Quality Index and the Epworth Sleepiness Scale global scores are available for 314 participants. Objective sleep measurements were collected over one week using a wrist Actiwatch (Philips Respironics, PA, USA) starting in 2017 and are available for 246 participants. Initially, actigraphy was proposed to all participants who underwent PET imaging, and was progressively offered to all interested participants, with up to 3 time points per participant now available. In parallel, the participants completed a sleep diary featuring daily information about their sleep/wake routine, including specific hours in bed (the sleep diaries have not been curated for global sharing). Actigraphy data was processed through the Actiware software (version 6.0) with a medium detection threshold for sensitivity of 40 activity counts/per min^24^. The data were collected using 15-second epochs. Time in bed detected by the Actiware algorithm was corrected when needed using information reported in the participants’ sleep diaries. For each sleep characteristic derived from complete nights of actigraphy, data provided include the day-to-day variability (standard deviation between actigraphy days) and the average over the week of actigraphy. Sleep characteristics include sleep duration (min), time in bed (min), sleep onset latency (min), sleep efficiency (%, sleep duration/time in bed), wake after sleep onset (min), and sleep fragmentation index (percent of mobile + immobile <1 min bouts over the sleep period representing restlessness, as defined by the Actiware algorithm).

### Neuroimaging data

#### MRI

MRI acquisition resumed in January 2019. Throughout the study, the same MRI scanner was used. The difference between the first and second Phase of data acquisition was that the scanner had been upgraded to a Prisma Fit and the 32-channel head coil was used exclusively. All scans were performed at the Cerebral Imaging Center of the Douglas Mental Health University Institute. Sequences acquired include 1mm^3^ 3D T1-weighted MPRAGE, 0.6mm^3^ 3D T2-weighted SPACE, 3mm^3^ resting-state multi-echo functional MRI, 1×1×3mm 3D FLAIR, and multi-shell diffusion imaging (**Table 1, supplementary Table 4**). These new MRI are available for 284 participants among which 206 participants underwent more than one MRI between ^25^19 and 2023 (after the upgrade, FLAIR data were only acquired in 2019-2020). One should consider that some harmonization or normalization steps may be needed if both Phase 1 and Phase 2 MRI data are used in the same analysis given that some sequences were slightly upgraded, but the single-site design of our study reduces confounding variability often present in larger studies. All MRIs are available in NIfTI file formats, organized according to the Brain Imaging Data Structure (BIDS) standard^28^. Anatomical images were defaced using a defacing algorithm shown to not significantly affect data processing outcomes^29^. This procedure is the same as the one applied to the Stage 1 MRI data, and every image was visually reviewed to ensure proper defacing. Single packages containing either all MRI or only anatomical images are available for download for this second phase of data acquisition.

#### PET

Newly available data also include Aβ (n=232) and tau (n=229) cross-sectional PET scans collected between 2017 and 2023. Aβ and tau PET scans were acquired on two consecutive days using a brain-dedicated Siemens/CTI high-resolution research tomograph (HRRT) at the McConnell Brain Imaging Centre of the Montreal Neurological Institute. This scanner provides high spatial resolution compared to most other scanners, with 2.4 mm at the center of the field of view. For Aβ, six 5-minute frames were acquired 40-70 minutes after the injection of 220 MBq (6 mCi) of [^18^F]NAV4694, whereas for tau four 5-minute frames were obtained 80-100 minutes after injecting an average of 370 MBq (10 mCi) of [^18^F]Flortaucipir^30^. We provide the raw PET images and the SUVR images in NIfTI for both tracers, organized according to the BIDS structure, which can all be downloaded in a zip file from the data repository. Qualitative control was done on the PET data to ensure that the MRI-PET registration was good.

#### MEG

Most of the participants that underwent a baseline PET scan between 2017 and 2019 also underwent a task-free MEG scanning session (n=114 are available in the Data Sharing Repository), which was obtained the same day as either the Aβ or the tau PET scan. MEG data were collected using a whole-head CTF MEG system (CTF MEG International Services LP, Coquitlam, BC, Canada) with 275 first-order axial gradiometer coils and 26 MEG reference sensors^31^. The raw MEG and T1-weighted MRI scans are available from the Open MEG Archive (OMEGA) repository^32^. Our objective is to eventually have them also available on the PREVENT-AD data repository.

## 4. ANALYTIC MEASURES

### Neuroimaging measures

Another innovation of the current release is the inclusion of analytic neuroimaging measures (or data derivatives) computed by imaging experts who preprocessed and analyzed the data and returned the numeric values to the StoP-AD Centre to be shared in CSV files **(Table 1)**.

#### MRI

The analytic measures include: 1) morphometric measurements of cortical thickness and brain volumes extracted from structural T1w-images in the Desikan-Killiany atlas using FreeSurfer^33^, 2) brain parcellations, and machine learning indices of aging and AD were extracted via the NiChart software (https://neuroimagingchart.com), 2) brain-wide functional connectivity matrices obtained across cortical regions of the Schaefer atlas (200 and 400 parcels)^34^ as well as within and between the 7 Yeo networks (i.e., visual, sensorimotor, dorsal attention, ventral attention, limbic, frontoparietal and default mode network) from resting-state timeseries of fMRI data preprocessed using fMRIPrep^35^, 3) diffusion tensor imaging (DTI) measures in 52 white matter bundles reconstructed from whole-brain tractograms computed from diffusion MRI (dMRI) data analyzed using the SCIL Tractoflow pipelines^36^, 4) white matter hyperintensity volumes in the whole brain and in each lobe as derived from T1w, T2w and FLAIR images that underwent a nonlinear automatic classification^37^ and 5) regional measures of cerebral blood flow in ml blood/100g tissue/min obtained from ASL sequences using BASIL from FSL and averaged over the regions of the Desikan-Killiany atlas.

#### PET

We now share standardized uptake value ratio (SUVR) pre-processed using a standard in-house developed pipeline (https://github.com/villeneuvelab/vlpp) that is based on the ADNI pipeline^30,33^. Briefly, Aβ- and tau-PET image frames were realigned, averaged and registered to the T1-weighted scan of each participant, which had been segmented according to the Desikan-Killiany atlas using FreeSurfer. SUVR images were obtained by dividing the signal uptake at each voxel by the average signal obtained from a reference region (i.e., the whole cerebellum for Aβ PET and the inferior cerebellum grey matter for tau-PET)^38,39^. We provide Aβ and tau PET SUVR values across the cortical regions of the Desikan-Killiany atlas. We also provide the global Aβ PET Centiloid values, which represent standardized data based on a 0-100 scale where 0 represents the average binding in young controls and 100 represents the average binding in patients with AD dementia^40^.

#### MEG

Time series were filtered to remove low-frequency drifts and DC-offset (high-pass set at 0.3Hz) and power-line noise (notch filter set at 60Hz and subsequent harmonics).^33^ Signal space projectors were computed to remove physiological artifacts related to cardiac activity and eye-blinks. Source imaging was applied to the preprocessed MEG data using dynamic statistical parametric mapping constrained to the cortical surface. Spectral power densities were computed for each 4-second epoch of the reconstructed time series from each vertex of the cortical surface. These were then averaged in the frequency domain to produce cortical maps of frequency-specific brain activity in the delta (2-4 Hz), theta (5-7 Hz), alpha (8-12 Hz), beta (13-30 Hz), gamma1 (30-60 Hz) and gamma2 (60-90 Hz) range, which were then normalized to the total spectral power to produce relative power estimates. Finally, these relative power estimates were averaged across the vertices comprising across the cortical regions of the Desikan-Killiany atlas parcellation. The analytical measures (regional relative spectral power estimates for each of the six frequencies) of the 98 participants’ data (out of 114 total) that passed quality control can be found in the Data Sharing Repository.

### Biofluid measures

#### Plasma and CSF biomarkers

Another important strength of this release is the incorporation of novel plasma markers including Aβ_1-40,_ Aβ_1-42_, glial fibrillary acidic protein (GFAP), neurofilament light (NfL), p-tau181, p-tau231 and p-tau217 that were part of an international collaboration with University of Gothenburg (**Table 3**). Plasma Aβ_1-40_, Aβ_1-42,_ GFAP, and NfL chain from 1350 (1257 shared) time points were analyzed and quantified using the commercial Neurology 4-plex E kit (503105; Quanterix)^30^. Plasma p-tau181 and p-tau231 from 1314 (1224 shared) time points were analyzed and quantified using in-house Single molecule array (Simoa) assays developed at University of Gothenburg^41,42^. Plasma p-tau217 concentration was measured from samples collected at three timepoints, and samples were analyzed using an in-house Simoa assay^43^ with overall repeatability and intermediate precision of < 10% for the biomarker measurement. Aβ_1-40_ and Aβ_1-42_ CSF measured using Lumipulse G automated immunoassay^44^ and CSF p-tau217 measured using the same method as plasma p-tau217^48^ are also available to researchers worldwide.

**Table 3.** Analytical fluid-derived measures in PREVENT-AD covering November 2011 to December 2023.

| Biofluid analytes | Participants<br>Internal<br>(Shared) | Visits<br>Internal<br>(Shared) | Assay | Investigator group |
| --- | --- | --- | --- | --- |
| **Cerebrospinal Fluid (CSF)** |  |  |  |  |
| Aβ<sub>1-42</sub> | 168 (161) | 496 (479) | ELISA (Innotest, Fujirebio) | J. Poirier |
| p-tau181, Total-tau | 169 (161) | 498 (480) | ELISA (Innotest, Fujirebio) | J. Poirier |
| ApoE | 104 (98) | 343 (357) | Luminex immunoassay (Milliplex APOMAG-62k, EMD-Millipore) | J. Poirier |
| PCSK9 | 100 (95) | 172 (164) | ELISA (Quantikine, R&D Systems) | J. Poirier |
| G-CSF, IL-15, IL-8 | 103 (98) | 335 (324) | Luminex immunoassay (Milliplex, EMD-Millipore) | J. Poirier |
| VEGF | 103 (98) | 310 (301) | Luminex immunoassay (Milliplex, EMD-Millipore) | J. Poirier |
| Aβ<sub>1-40</sub>, Aβ<sub>1-42</sub> | 103 (98) | 116 (110) | Automated chemiluminescent immunoassay (Lumipulse G, Fujirebio) | K. Blennow, H. Zetterberg |
| p-tau217 | 183 (174) | 203 (193) | In-house Simoa assay developed at University of Gothenburg | K. Blennow, H. Zetterberg |
| Proteomics (SomaScan 7K) | 69 (67) | 69 (67) | High-throughput aptamer-based proteomics assay (SomaScan 7K, SomaLogic) | J. Poirier |
| **Plasma** |  |  |  |  |
| Aβ<sub>1-40</sub>, Aβ<sub>1-42</sub>, GFAP, NfL | 373 (338) | 1350 (1257) | Simoa assay (Neurology 4-plex E, lot 503105, Quanterix) | K. Blennow, H. Zetterberg |
| p-tau181, p-tau231 | 369 (336†) | 1314 (1224) | In-house Simoa assay developed at University of Gothenburg | K. Blennow, H. Zetterberg |
| p-tau217 | 382 (346) | 952 (877) | In-house Simoa assay developed at University of Gothenburg | K. Blennow, H. Zetterberg |

†n=335 for p-tau181
*Abbreviations:* Amyloid-beta (Aβ ), Apolipoprotein E (ApoE), enzyme-linked immunosorbent assay (ELISA), glial fibrillary acidic protein (GFAP), granulocyte colony stimulating factor (G-CSF), interleukin (IL), neurofilament light (NfL), phosphorylated (p), proprotein convertase subtilisin/kexin type 9 (PCSK9), single molecular array (Simoa), vascular endothelial growth factor (VEGF)

#### CSF proteomics

The SOMAscan 7k panel measured 7288 aptamers mapping to approximately 6600 unique protein targets in the baseline CSF sample of 67 participants from the PREVENT-AD cohort are shared. Protein measurements are reported in relative fluorescence unit (RFU). Initial data normalization procedures were performed by SomaLogic. Briefly, normalization was performed at the sample level. Aptamers were then divided into three normalization groups: S1, S2 and S3; based on the observed signal to noise ratio in technical replicates and samples. This division was done to avoid combining features with different levels of protein signal for additional normalization steps. Median normalization was then performed to remove other assay biases such as protein concentration, pipetting variation, variation in reagent concentrations, and assay timing among others^45^. Finally, normalization to a reference was performed on individual samples to account for additional technical variance as well as biological variance. This normalization step was performed using iterative Adaptive Normalization by Maximum Likelihood, a modification of median normalization, until convergence was reached. Additional details on normalization procedures are documented in SomaLogic’s technical note^46^.

### Genetics

In addition to providing *APOE* genotype, we are now providing the risk/protective polymorphisms of the 74 genes that have been formally associated with late-onset AD in the large GWAS analyses performed by Bellenguez et al. in 2022^47^. Automated DNA extraction from buffy coat samples was performed using the QIAsymphony DNA mini kit (Qiagen, Hilden, Germany). Genotyping was performed using the Omni2.5-8 BeadChip (Illumina, San Diego, CA, USA). PLINK (http://pngu.mgh.harvard.edu/purcell/plink/) was used to filter gender mismatches, filter missingness at sample-level (< 5%) and SNP-level (< 5%), assess sample heterozygosity and filter SNPs in Hardy-Weinberg disequilibrium (p>0.001). Only post-imputed SNPs with an info score > 0.7 were kept^48^. These data are available for 294 participants.

## 5. BIOMARKERS AND CLINICAL TRAJECTORIES OF PREVENT-AD PARTICIPANTS

The PREVENT-AD cohort has enabled many studies that augmented our understanding of the preclinical phase of the disease as well as its prevention. These studies have also highlighted the complexities of disease progression and validate the use of biomarkers to identify individuals who have entered the preclinical phase of the disease. As of January 2025, at least 86 scientific publications have featured data from PREVENT-AD, although this number may be larger considering that we originally did not ask open users to add “for the PREVENT-AD Research Group” in the author line of their publications, making the publications using the open datasets difficult to track.

A focus of the phase 2 of data acquisition is the in-vivo measures of Aβ and tau using both fluid and imaging biomarkers. In 2022, two multi-cohort longitudinal studies, including one using PREVENT-AD data, showed that half of CU individuals identified as being abnormal on both Aβ (A+) and tau (T+) PET biomarkers developed MCI or dementia when followed for a mean of 3.5 years^20,49^. Individuals only positive on the Aβ scan (A+T-) were not at increased risk of developing MCI or dementia when compared to A-T-individuals. Using additional follow-up data collected in the phase 2 of data acquisition, we found that with 2.4 years longer of follow-up, 100% of PREVENT-AD participants who were A+T+ in this first publication^20^ progressed to MCI or dementia during the additional follow-up^50^ (**Figure 4**). Importantly, in the A+T-group, the percentage of progressors to MCI went from 9% to 42% with additional follow-up, suggesting that many A+T-PET individuals are on the path of AD dementia and probably accumulate tau tangles over time, even if they do not yet show high uptake on tau PET scans early on^51^. Similar results were found when using CSF and plasma p-tau217, as most PREVENT-AD participants with abnormal p-tau217 values progressed from CU to MCI within a 10-year window^50^. These results are concrete examples of the richness of the PREVENT-AD follow-ups as well as the predictive value of the biomarkers used. They also stress the fact that AD is a progressive disease, and that the length of follow-up can dramatically change the interpretation of the results.

**Figure 4.**
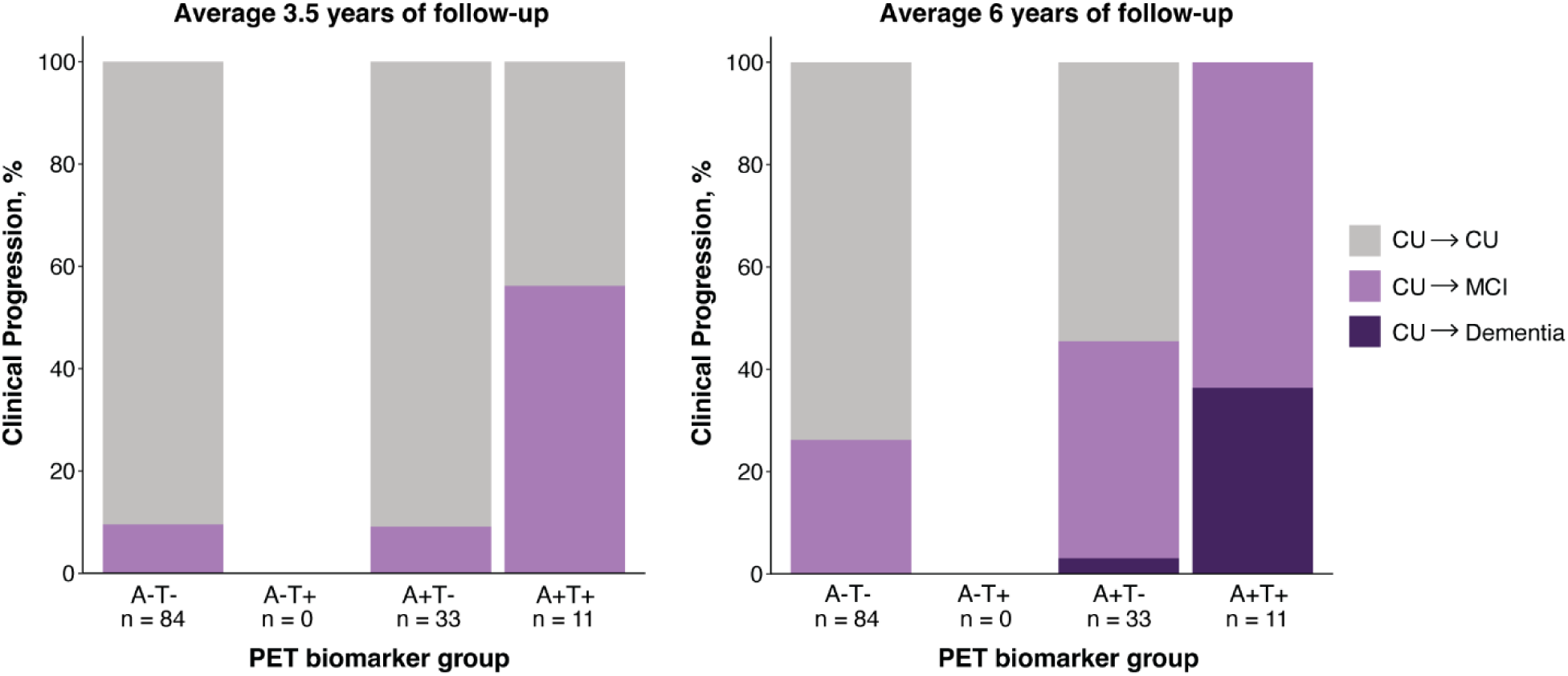
Clinical status profiles and progression in the PREVENT-AD cohort. Percentage of clinical progression (mild cognitive impairment (MCI) or dementia) across Aβ and tau PET biomarker profiles. The first graph is data as reported in Strikwerda-Brown et al (2022), *JAMA Neurology* (left) and the second is after an additional 2.4 years of follow-up (right) (n=128). *Abbreviations*: CU: cognitively unimpaired older adults; MCI: mild cognitive impairment; A+T+: Aβ positive/tau positive; A+T-: Aβ positive/tau negative; A-T+: Aβ negative/tau positive; A-T-: Aβ negative/tau negative

In pursuit of the StoP-AD Centre’s aim to find ways to delay the onset of AD, we added many questionnaires in 2016 to assess the impact of lifestyle factors, behavior and personality traits, alone or in combination, on AD risk and progression, with the ultimate goal to provide empirical evidence that could support new preventive interventions. Lower education^22,52^, sleep characteristics^24,25^, untreated vascular diseases^53^ high neuroticism^22^, high perseverative negative thinking^23^ and lower mindfulness traits^21^ are all factors that have been associated with Aβ and/or tau in the PREVENT-AD study. Behavior and lifestyle factors in midlife and late-life, such as physical activity, cognitive activity, healthy diet, and social activity engagement have also been associated with brain integrity and cognitive trajectories^26,27,54–57^, stressing the need for multimodal preventive interventions.

One novelty of Phase 2 is the addition of MEG, a neuroimaging modality that is rarely acquired in large longitudinal AD studies. We used the MEG data across multiple projects to investigate the relationship between neurophysiological alterations, AD pathology, and neurotransmitter systems ^58–61^. Our findings show that early Aβ accumulation accelerates cortical neurophysiology, while the presence of medial temporal tau pathology shifts activity toward slower oscillatory states, which are associated with cognitive decline^31^. We further identified that these neurophysiological changes align topographically with key neurotransmitter systems, particularly cholinergic, serotonergic, and dopaminergic pathways, and that Aβ preferentially deposits along neurochemical boundaries^62^. These results provide insight into the interplay between neurophysiology, pathology, and neurochemistry in preclinical AD, with implications for early detection and pharmacotherapeutic strategies.

The focus on genetics and fluid biomarkers also allowed us to explore biological factors that confer neuroprotection against AD pathology and brain degeneration^63^, and biological factors that act as disruptors of brain integrity in the preclinical phase of the disease^64^. The longitudinal nature of this cohort and its agglomeration with other datasets also allowed us to uncover complex and non-linear relationships that would not be possible using cross-sectional data.

Contactin 5 for instance, a neuronal membrane protein involved in key processes of neurodevelopment and terminal remodeling, was found to increase progressively in CU individuals before decreasing in individuals with cognitive impairment^48^.

PREVENT-AD data have also served the field of neuroscience more broadly. Open MRI data have contributed to large neuroscience initiatives such as the creation of a lifespan brain chart that was derived from 101,457 individuals^8^ and the creation of five dominant patterns of brain atrophy derived from 49,482 individuals^65^. Behavioral data were used to validate a prognostic risk score for development and spread of chronic pain^66^. In a multicohort initiative, it was also found that *APOE* ε2 protected against cognitive decline in men, but not in women^67^. These are just some examples of the impact of this rich multimodal longitudinal dataset.

## FUTURE DIRECTIONS

Recruitment re-opened in 2022, and we now aim at testing more preventive interventions. Dr. Maiya Geddes is currently leading a preventive intergenerational randomized controlled trial to enhance physical activity and slow cognitive decline^68^. We also received funding to conduct a clinical trial testing the possible protective impact of a dual orexin receptor antagonist medication on Aβ and tau plasma levels. We are aware that these interventions could compromise longitudinal follow-ups by modifying disease course, but believe that if we succeed at modifying disease trajectories, the gain will largely overcome the loss of the observational follow-ups.

Dr. Nathan Spreng, Ph.D., also collected a large amount of new behavioral and MRI data (NIA; R01AG068563; 3R01AG068563-04S1) including the Mnemonic Similarity Task pattern separation task that has been associated with iron deposition and distribution across the hippocampus among PREVENT-AD participants^69^. The Mnemonic Similarity Task and multi-shell diffusion sequences have also led to key insights into brain microstructural alterations associated with AD biomarkers, *APOE4,* and cognition^70–72^. Additional PET, MEG and behavioral data have also been collected as part of other collaborator projects. We are currently curating these data for future releases. Moving forward, we are also asking users who derive analytic measures to send them to us with methods documentation so they can be integrated in the Data Sharing Repository. This simple way of giving back will improve our repository and expand the analytic possibilities while also promoting users’ work. A concrete example of this reciprocal sharing strategy is the five dominant patterns of brain atrophy^65^ quantified by an independent group of investigators that have been shared back and are now available to users.

A subset of participants have consented to brain donation, and postmortem brain specimens will be transferred to the Douglas-Bell Canada Brain Bank following a standardized procedure^73^After reception, hemispheres are separated and one unsliced hemisphere (right or left, in alternation) is fixed and scanned to provide an end-stage MRI-based estimate of disease severity.

Neuropathology assessment is then performed to provide confirmation of AD diagnosis and assess the presence of co-pathology. The protocols are in place, but no brains have been received yet. These ex-vivo data will allow us to validate our in-vivo biomarkers and assess research questions that would not be possible using current in-vivo markers such as assessing the comorbid impact of TDP-43^74^.

The main limitation of the PREVENT-AD cohort is its low levels of racial, ethnic and socioeconomic diversity. We know that AD risk varies between represented and underrepresented populations and that this variation might be due to biological, lifestyle and socioeconomic factors (see^75^). To develop effective preventive AD treatments, we need large, diverse cohorts to understand these differences. PREVENT-AD, when taken alone, is unfortunately not a diverse cohort, but we hope that by sharing data that will be pooled with other datasets, we will help understand genetics and lifestyle diversity. Experts in health equity research are also welcome to use the StoP-AD Centre’s infrastructure or more adapted infrastructure to help us increase diversity.

## CONCLUSION

The PREVENT-AD is a longitudinal cohort of individuals with a first-degree family history of sporadic AD from the greater Montreal area in Canada, who are deeply phenotyped and have more than 10 years of follow-up data. Multimodal biomarker assessment (MRI, PET, CSF, plasma) and lifestyle data is available for all participants. The data of participants who agreed to sharing have also been made available to researchers worldwide. While sharing data with researchers worldwide takes time and funding, we believe it is absolutely needed to move the field forward and find ways to stop AD.

## USAGE NOTES

The data on participants who agreed to sharing can be accessed via the PREVENT-AD Data Sharing Repository https://registeredpreventad.loris.ca/. Given that sensitive information is shared, access is restricted to qualified researchers or physicians. The PREVENT-AD coordinator will verify information of all principal investigators requesting an account before granting access, after which data can be shared with trainees that are under their supervision and for whom they take the full responsibility of data usage. All terms of use must be agreed to when requesting an account, at https://registeredpreventad.loris.ca/login/request-account/. In brief, users must not attempt to sell or claim intellectual property rights in the PREVENT-AD dataset, users must obtain any needed ethics approvals for their use of the PREVENT-AD dataset, the data must be used for ‘neuroscience research’, the data must not be redistributed, and any attempt to re-identify participants is forbidden. When used in publications, the methods section must state PREVENT-AD as the source of the data and the manuscript describing the first open release^4^ or the current release must be cited depending on the data used.

The PET, MEG, and actigraphy data along with the behavioral factors assessed with questionnaires were not administered at a specific annual visit as the other tests provided. We thus provided the closest annual visit label and/or the date of acquisition (month and year) to help with data usage.

Lastly, we have created data dictionaries and detailed methods as well as documentation on how to access the data so users can be autonomous when using the data. With the sunset of the Brain Canada Platform grant (2022-2025), we will no longer have dedicated staff or funding to support users working with the data. In this new release, we also provide analytic measures to ease data usage by non-neuroimaging specialists.

## Data Availability

All data produced in the present work are available online for researchers at:
https://registeredpreventad.loris.ca/

## CONSENT STATEMENT

All participants provided written informed consent.

## ACKNOWLEDGEMENTS

The authors wish to acknowledge the staff of the PREVENT-AD as well as of the Cerebral Imaging Centre of the Douglas Mental Health University Institute and of the PET unit of the McConnell Brain Imaging Centre of the Montreal Neurological Institute. A full listing of the PREVENT-AD Research Group members can be found at https://preventad.loris.ca/acknowledgements/acknowledgements.php?date=[2023-01-01]. We also thank the participants of the PREVENT-AD cohort for dedicating their time and energy to helping us collect these data and agreeing to share their data openly. We would like to acknowledge Bill Jagust, Suzanne Baker, and Susan Landau who shared with us coded to helped us build our PET processing pipeline and Ilana R. Leppert who did the pulse sequence programming required for the MPM protocol.

