## Supplemental Table for "The PREVENT-AD cohort: accelerating Alzheimer’s disease research and treatment in Canada and beyond"

**Supplementary Table 1. Inclusion and Exclusion Criteria.**

**Inclusion criteria**

-Self-reported parental or multiple-sibling (2 or more*) history of Alzheimer-like dementia

-Age 60 years or older (persons aged 55–59 years and < 15 years younger than their affected index relative were also eligible)

-Minimum of 6 years of formal education

-Study partner available to provide information on cognitive status

-Sufficient fluency in spoken and written French and/or English

-Ability and intention to participate in regular visits

-Agreement for periodic donation of blood and urine samples

-Agreement to participate in periodic multimodal assessments via MRI and LP for CSF collection (LP optional)

-Agreement to limit use of medicines as required by clinical trial protocols, if applicable

-Provision of informed consent of the different protocols

**Exclusion criteria**

-Cognitive disorders - Known or identified during eligibility assessments (MoCA and CDR or exhaustive neuropsychological evaluation when needed)

-Use of acetyl-cholinesterase inhibitors including tacrine, donepezil, rivastigmine, galantamine

-Use of memantine or other approved prescription cognitive enhancer

-Use of vitamin E at>600 i.u. / day or aspirin at > 325 mg / day

-Use of opiates (oxycodone, hydrocodone, tramadol, meperidine, hydromorphone)

-Use of NSAIDs or regular use of systemic or inhalation corticosteroids

-Clinically significant hypertension (accepted if controlled medically), anemia, significant liver or -kidney disease

-Concurrent use of warfarin, ticlopidine, clopidrogel, or similar anti-coagulant

-Current plasma Creatinine > 1.5 mg/dl (132 mmol/l)

-Current alcohol, barbiturate or benzodiazepine abuse/dependence

*Notes:**8 participants had only 1 sibling affected with AD-like dementia.

*Abbreviations:* MRI: magnetic resonance imaging; LP: lumbar puncture; CSF: cerebrospinal fluid; MoCA: Montreal Cognitive Assessment; CDR: Clinical Dementia Rating; NSAID: non-steroidal anti-inflammatory drug.

Adapted from Tremblay-Mercier J, Madjar C, Das S, et al. Open science datasets from PREVENT-AD, a longitudinal cohort of pre-symptomatic Alzheimer's disease. *Neuroimage Clin*. 2021;31:102733. doi:10.1016/j.nicl.2021.102733

**Supplementary Table 2 Demographics**

| **Variable** | **Level** | **Internal**  **(n=387)** | **Shared**  **(n=348)** |
| --- | --- | --- | --- |
| Age | mean (sd) [min – max] | 63.2 (5.1) [54.9 – 83.3] | 63.2 (5) [54.9 – 83.2] |
|  | Female | 277 (71.6) | 249 (71.5) |
|  | Male | 110 (28.4) | 99 (28.4) |
| Ethnicity | White | 381 (98.4) | 344 (98.9) |
|  | other | 6 (1.6) | 4 (1.1) |
| Handedness^+^ | Ambidextrous | 17 (4.4) | 16 (4.6) |
|  | Left-handed | 22 (5.7) | 18 (5.2) |
|  | Right-handed | 346 (89.9) | 312 (90.2) |
| Education | mean (sd) [min – max] | 15.4 (3.4) [7 – 29] | 15.4 (3.3) [7 – 29] |
| MoCA^†^ | mean (sd) [min – max] | 28 (1.6) [23 – 30] | 28 (1.6) [23 – 30] |
| *APOE*^+^ | *APOE4* non-carrier | 237 (61.6) | 213 (61.2) |
|  | *APOE4* carrier | 148 (38.4) | 135 (38.8) |

*Notes:* ^†^Data missing for 1 participant. ^+^Data missing for 2 participants (for *APOE* the data are only missing in the internal dataset).

| **Supplementary Table 3. PREVENT-AD MRI parameters Phase 1.** | | | | |
| --- | --- | --- | --- | --- |
| **Scan type** | **Sequence** | **Acquisition parameters** | **Resolution (mm^3^)** | **Scan time (min)** |
| T1-weighted anatomical | MPRAGE | 3D sagittal; TR = 2300 ms; TE = 2.98 ms; TI = 900 ms; a = 9°; FOV = 256x240x176 mm; phase encode A-P; BW = 240 Hz/px; GRAPPA 2. | 1x1x1 | 5.12 |
| Fluid attenuated T2-weighted image | FLAIR | 3D sagittal; TR = 5000 ms; TE = 388 ms; TI = 1800 ms; FOV = 256x256x176 mm; phase encode A-P; BW = 781 Hz/px; GRAPPA 2. | 1x1x1 | 6.27 |
| T2*-weighted anatomical | GRE | 3D transversal; TR = 650 ms; TE = 20 ms; a = 20°; FOV = 350x263x350 mm; phase encode R-L; BW = 200 Hz/px. | 0.8x0.8x2 | 5.34 |
| Multi-echo T2*-weighted anatomical | Multi-echo GRE | 3D transversal; TR = 44 ms; TE = [2.84, 6.2, 9.56, 12.92, 16.28, 19.64, 23, 26.36, 29.72, 33.08, 36.44, 39.8]ms; a = 15°; FOV = 350x263x350 mm; phase encode R-L; BW = 500 Hz/px. | 1x1x1 | 9.44 |
| High-resolution T2-weighted anatomical | T2-weighted SPACE | 3D coronal; TR = 2500 ms; TE = 198 ms; FOV = 350x263x350 mm; phase encode R-L; GRAPPA = 2; BW = 710 Hz/px. | 0.6x0.6x0.6 | 10.02 |
| T1 map | MP2RAGE | 3D sagittal; TR = 5000 ms; TE = 2.91 ms; TI = [700,2500]ms; a = [4°, 5°]; FOV = 256x240x176 mm; phase encode A-P; BW = 240 Hz/px; GRAPPA 2. | 1x1x1 | 8.22 |
| Diffusion-weighted imaging (DWI) | EPI | 2D transversal; TR = 9300 ms; TE = 92 ms; FOV = 192x192x130 mm; phase encode A-P; BW = 1628 Hz/px.  b = [0,1000] s/mm2 with 1, 64() directions | 2x2x2 | 10.15 |
| Perfusion imaging | Pseudo continuous-ASL (PCASL) EPI | TR = 4000 ms; TR = 10 ms; a = 90°; FOV = 256x256mm; 16 slices; phase encode A-P; BW = 3004 Hz/px; GRAPPA 2; phase PF 7/8.  Label offset = 100 mm; post label delay = 900 ms. | 4x4x7 | 5.32 |
| resting-state functional MRI (fMRI) | EPI | 2D axial; TR = 2000 ms; TE = 30 ms; a = 90°; FOV = 256x256 mm; 32 slices; phase encode A-P; BW = 2442/px. | 4x4x4 | 5.04 |
| Task functional MRI (fMRI) (Encoding and Retrieval) | EPI | 2D axial; TR = 2000 ms; TE = 30 ms; a = 90°; FOV = 256x256 mm; 32 slices; phase encode A-P; BW = 2442/px. | 4x4x4 | 6.10; 15.10 |
| The high spatial resolution of the T2 sequence was for hippocampal subfield related imaging. Table adapted from Tremblay-Mercier J, Madjar C, Das S, et al. Open science datasets from PREVENT-AD, a longitudinal cohort of pre-symptomatic Alzheimer's disease. *Neuroimage Clin*. 2021;31:102733. doi:10.1016/j.nicl.2021.102733  *Abbrevations:* TR = repetition time; TE = echo time; TI = inversion time; FOV = field of view; MPRAGE = magnetization prepared gradient echo; FLAIR = fluid attenuated inversion recovery; PCASL = pseudo-continuous arterial spin labeling | | | | |

**Supplementary Table 4. PREVENT-AD MRI parameters Phase 2.**

| **Scan type** | **Sequence** | **Acquisition parameters** | **Resolution (mm3)** | **Scan time (min)** |
| --- | --- | --- | --- | --- |
| T1-weighted anatomical | MPRAGE | 3D sagittal; TR = 2300ms; TE = 2.96ms; TI = 900ms; a = 9°; FOV = 256x256x192 mm; phase encode A-P; BW = 240Hz/px; GRAPPA 2. | 1x1x1 | 5.30 |
| Resting-state functional MRI (fMRI) | Multi-echo gradient-echo EPI | 2D axial; TR = 1000ms; TE's = 12, 30.11, 48.22ms; a = 50°; FOV = 240x240 mm; 48 interleaved descending slices; phase encode A-P; BW = 2500Hz/px; GRAPPA 2; multi-band acceleration = 4; phase PF 7/8.  6 images acquired with phase encode A-P and P-A for distortion correction.  TR = 4041ms; TE = 48ms; SMS 1. | 3x3x3 | 10.24;  AP, PA=0.28 |
| High-resolution T2-weighted anatomical | SPACE | 3D coronal; TR = 2500ms; TE = 198ms; turbo factor = 143; FOV = 206x206x205 mm; phase encode R-L; BW = 625Hz/px; CAIPIRINHA 2x2; PF 6/8. | 0.64x0.64x0.64 | 7.35 |
| Multi-shell diffusion-weighted imaging (DWI) | Pulse gradient spin echo (PGSE) EPI | 2D axial; TR = 3000ms; TE = 66 ms; a = 90°; FOV = 220x220 mm; 81 slices; phase encode P-A; BW = 2272Hz/px; phase PF 7/8; GRAPPA 2; SMS 3.  b = [0, 300, 1000, 2000] s/mm^2^ with [9, 7, 29 64] directions  5 b=0 images with phase encode A-P acquired for distortion correction. | 2x2x2 | 5.49;  b0=0.35 |
| Fluid-attenuated T2-weighted image^1^ | FLAIR | 3D axial; TR = 6000ms; TE = 356ms; TI = 2200ms; a = 90°; FOV = 256x232 mm; 60 slices per slab; phase encode A-P; BW = 781Hz/px; GRAPPA 2; 1.4 averages. | 1x1x3 | 3.50 |

The high spatial resolution of the T2 sequence was for hippocampal subfield related imaging.

*Abbrevations:* TR = repetition time; TE = echo time; TI = inversion time; FOV = field of view; MPRAGE = magnetization prepared gradient echo; FLAIR = fluid attenuated inversion recovery; PCASL = pseudo-continuous arterial spin labeling
